# Validation of Synthesa AI, a Large Language Model-Based Screening Tool for Systematic Reviews: Results from Nine Studies

**DOI:** 10.1101/2025.07.16.25331632

**Authors:** Lefteris Teperikidis PharmD, Christos Trampoukis, Kyiakos Polymenakos

**Affiliations:** Synthesa, Inc., 19 West 24th St., New York, NY 10010, USA; Clinical Research Unit, Special Unit for Biomedical Research and Education (SUBRE), School of Medicine, Aristotle University of Thessaloniki, Thessaloniki, Greece; Third Department of Cardiology, Ippokratio General Hospital, Aristotle University of Thessaloniki, Thessaloniki, Greece

## Abstract

Systematic review screening is often burdensome, prone to human error, and requires significant manual effort. Synthesa AI, a large language model (LLM)-based tool, was developed to address these challenges by offering a transparent and prompt-driven approach to abstract screening. In this validation study, Synthesa AI was evaluated across 17 benchmark meta-analyses encompassing nine clinical domains. Using user-defined PICOS criteria, the tool screened a total of 270,626 abstracts retrieved from PubMed and Scopus. Synthesa AI accurately identified all 163 benchmark-included studies, yielding a sensitivity of 100% and a pooled specificity of 99.4%. Remarkably, it reduced reviewer workload by 91.7%, flagging only 1,797 abstracts for manual review. Furthermore, the tool identified 32 relevant studies that had been missed in the original reviews, representing a 19.6% increase in evidence yield. These findings demonstrate that Synthesa AI delivers high precision, efficiency, and reproducibility in systematic review workflows. Its auditable and deterministic architecture adheres to Good Machine Learning Practice (GMLP) guidelines, making it suitable for both academic and regulatory applications. Synthesa AI represents a promising solution for living systematic reviews and large-scale evidence synthesis initiatives, offering a transformative alternative to traditional human-led screening.

## Introduction

Systematic reviews and meta-analyses constitute the methodological cornerstone of evidence-based medicine, serving as essential instruments for aggregating clinical evidence, informing practice guidelines, and underpinning regulatory and policy decisions (1–3). Their value in synthesizing data from randomized controlled trials and observational studies is well-established across practically every domain in Medicine. However, the initial stages of conducting a systematic review—particularly the screening of titles and abstracts—remain notoriously inefficient. These phases are not only labor-intensive and time-consuming but also susceptible to inter-reviewer variability, leading to inconsistencies in the selection of eligible studies (4–6). The limitations of current human-centric workflows are especially pronounced in high-throughput or rapidly evolving research landscapes, where evidence accumulates at a pace that outstrips the practical capacities of human teams (7–9).

The growing volume of biomedical literature has created a methodological bottleneck that threatens the timeliness, reproducibility, and scalability of evidence synthesis (10, 11). In fields such as cardiovascular medicine, obesity, and diabetes—where systematic reviews may necessitate the appraisal of thousands of citations—screening processes often consume disproportionate resources while risking the inadvertent exclusion of relevant studies. Conventional machine learning approaches to semi-automated screening have demonstrated incremental improvements but are constrained by rigid training pipelines and limited generalizability across clinical contexts (5, 12, 13). The recent advent of large language models (LLMs) introduces a transformative opportunity to reconfigure the screening paradigm. These models, grounded in transformer architectures and pretrained on vast corpora of biomedical and general-domain text, have demonstrated remarkable aptitude for natural language understanding, contextual reasoning, and decision support—attributes that are well-aligned with the interpretive demands of systematic review screening tasks (14, 15).

Synthesa AI is a novel abstract screening platform that operationalizes these capabilities within a reproducible and customizable framework for evidence synthesis. Built upon state-of-the-art LLMs, Synthesa AI emulates the inferential logic applied by human reviewers by evaluating biomedical abstracts against user-defined inclusion criteria structured according to the PICOS framework (Population, Intervention, Comparator, Outcome, and Study Design). The system is designed to produce transparent, auditable, and reproducible decisions, flagging abstracts as “Included,” “Excluded,” or “Potentially Relevant,” with accompanying justifications. Importantly, the tool does not rely on pre-labeled training data and instead uses deterministic prompts to drive decision-making, ensuring consistent outputs across screening runs for a given input and prompt configuration.

The present report introduces Synthesa AI to the academic and regulatory communities by providing results from nine independent validation studies conducted across a diverse array of clinical domains. These studies were conceived to evaluate the tool’s operating characteristics—specifically sensitivity, specificity, speed, reproducibility, and scalability—relative to human review benchmarks. In each validation, Synthesa AI was assessed for its ability to accurately identify all studies included in published systematic reviews or meta-analyses, while also evaluating its capacity to uncover additional relevant studies that had been overlooked by prior human screening efforts. The validation design also considered regulatory acceptability, aligning the development of Synthesa AI with emerging frameworks such as the U.S. Food and Drug Administration’s Good Machine Learning Practice (GMLP) principles.

## Methods

### Study Overview

This study evaluated the performance of Synthesa AI, a large language model-based abstract screening tool, across nine independent validation exercises. Each validation study was designed to assess the tool’s sensitivity, specificity, and overall screening efficiency in identifying eligible studies for systematic reviews. The validation paradigm was constructed to simulate real-world evidence synthesis workflows and to test the model’s generalizability across a wide range of clinical questions, therapeutic domains, and study populations.

Eight of the nine validation studies were retrospective in design and benchmarked against published meta-analyses from high-impact, peer-reviewed journals, including one Cochrane systematic review. These benchmark reviews were selected on the basis of methodological rigor, publication recency, and relevance to high-throughput areas of clinical research, including cardiovascular medicine, critical care, psychiatry, respiratory diseases, infectious diseases, and immunization. The remaining validation study—the ninth—combined evidence from several previously published systematic reviews with a prospective, double-screened, blinded human adjudication process, enabling a hybrid benchmark that incorporates both historical and real-time human gold standards.

### Synthesa AI Architecture and Screening Protocol

Synthesa AI is a deterministic LLM-powered tool engineered for binary relevance classification of biomedical abstracts based on a structured PICOS (Population, Intervention, Comparator, Outcome, Study Design) framework. For each validation study, the user (i.e., the analyst team) provided a PICOS prompt derived from the eligibility criteria of the original meta-analysis. Once provided with the user-defined PICOS, the tool applies a two-stage screening logic: an initial filter to exclude studies based on publication type and study design (e.g., case reports, editorials, or irrelevant study types), followed by PICO element extraction and comparison against user-defined criteria.

Each abstract is processed independently and classified as “Included,” “Excluded,” or “Potentially Relevant.” These classifications are supplemented with accompanying rationales and flagged terms to facilitate downstream auditability and human review. Synthesa AI operates in a stateless fashion, ensuring reproducibility of outputs for a given set of inputs and prompts.

### Validation Workflow

The benchmark studies were used to calibrate the tool prior to full-scale screening. Calibration involved the extraction of all abstracts from studies included in each benchmark meta-analysis. These abstracts were run through Synthesa AI using preliminary PICOS prompts. In cases where the tool failed to identify one or more of the included studies, the prompt was iteratively refined until all included studies were successfully flagged as “Included” or “Potentially Relevant.” This calibration process was essential to ensure that the tool was sufficiently sensitive to detect all known eligible studies, and it mirrors the pragmatic tuning step employed in actual systematic review workflows.

Following calibration, a new, broader literature search was conducted for each clinical topic using PubMed and Scopus, compared to that used in the benchmark reports. The goal was to execute a more inclusive and sensitivity-maximizing search strategy than that employed in the original meta-analyses. Synthesa AI was then used to screen the entire corpus of retrieved abstracts. In all cases, flagged abstracts underwent blinded human review to adjudicate final inclusion status.

### Human Adjudication and Gold Standard Comparison

For the eight benchmark-driven validation studies, the reference standard was the list of studies included in the published meta-analyses. Sensitivity was defined as the proportion of these benchmark-included studies that were correctly flagged as “Included” or “Potentially Relevant” by Synthesa AI. Specificity was calculated as the proportion of excluded abstracts that were correctly classified as “Excluded.”

The ninth validation study used a hybrid gold standard. In this case, all flagged abstracts were subjected to prospective double human review, with discrepancies resolved by consensus. This allowed a direct comparison between Synthesa AI’s screening decisions and those of blinded human reviewers. The inclusion of this prospective adjudication process allowed us to evaluate the tool’s performance in a setting without reliance on prior publications, thereby addressing potential biases introduced by calibration to known included studies.

### Data Synthesis and Statistical Analysis

Across all nine studies, pooled sensitivity and specificity estimates were calculated. Descriptive statistics were used to quantify the number of abstracts flagged, the number of relevant studies identified, and the proportion of newly identified studies not captured by benchmark reports. Screening burden reduction was estimated as the percentage decrease in the number of abstracts requiring human review, relative to full manual screening.

## Results

### Study 1: STEMI and Multivessel Percutaneous Coronary Intervention (PCI) Strategies

The first validation study focused on the identification of RCTs comparing revascularization strategies in patients with ST-elevation myocardial infarction (STEMI) and multivessel coronary artery disease. We included a prospective double human screening, while the benchmark comprised six published meta-analyses, including a total of 24 unique included studies (16–21). Synthesa AI and the two human reviewers screened a total of 5,460 abstracts. The tool flagged 188 abstracts for human review. Within this flagged subset, all 24 benchmark-included RCTs were correctly classified as relevant, yielding a sensitivity of 100%. Moreover, Synthesa AI identified two additional eligible RCTs that had not been captured by the benchmark reviews—one that had been published after the benchmarks’ respective search windows, and one that had been published prior (22, 23). These two studies were also flagged as relevant by both human reviewers. Ultimately, of the 188 flagged citations, 162 were ultimately judged not to meet inclusion criteria, resulting in a specificity of 97.2%.

### Study 2: Inhaled Reliever Therapies for Asthma

The second validation study evaluated Synthesa AI in the context of reliever inhaler therapies for asthma. The benchmark for this analysis was the network meta-analysis conducted by Rayner et al. (24), which had included 23 RCTs indexed in PubMed. In contrast to the benchmark study’s screening of 3,179 abstracts, a broader PubMed query conducted for this validation yielded 30,719 abstracts. Synthesa AI screened the entire corpus and flagged 247 abstracts for manual review. The tool successfully identified all 23 PubMed-indexed RCTs reported in the Rayner et al. meta-analysis, achieving a sensitivity of 100%. In addition, Synthesa AI identified 15 further eligible RCTs that were not included in the benchmark analysis (25–39), expanding the total number of included studies to 38 and increasing the overall sample size from 45,117 to 64,036 participants. Of the 247 flagged citations, 209 were adjudicated as false positives, yielding a specificity of 99.2%.

### Study 3: Antidepressants for Irritable Bowel Syndrome (IBS)

The third validation study aimed to replicate and extend the findings of a recent meta-analysis conducted by Temido et al. (40), which evaluated the efficacy of antidepressants in the treatment of irritable bowel syndrome (IBS). A literature search using both PubMed and Scopus yielded a total of 28,645 abstracts. Synthesa AI screened this full dataset and flagged 86 abstracts for human adjudication. The tool successfully identified all 20 RCTs included in the benchmark meta-analysis, resulting in a sensitivity of 100%. Furthermore, it identified six additional RCTs that reported binary outcomes suitable for meta-analysis but had not been included in the benchmark publication (41–45). An additional two RCTs were also detected that, while relevant, did not contain extractable outcome data compatible with quantitative synthesis (46, 47). These findings brought the total number of eligible studies to 28. Of the 86 abstracts flagged by Synthesa AI, 58 were ultimately excluded, resulting in a specificity of 99.38%.

### Study 4: Antiplatelet versus Anticoagulation Therapy in Heart Failure with Sinus Rhythm

The fourth validation study evaluated Synthesa AI’s ability to replicate the findings of a Cochrane systematic review that compared the efficacy and safety of antiplatelet agents versus anticoagulants in patients with heart failure who maintained sinus rhythm (48). A PubMed search yielded a total of 48,250 abstracts, which were then screened by Synthesa AI. The tool flagged 103 abstracts for human review and correctly identified all four RCTs included in the benchmark Cochrane review, resulting in a sensitivity of 100%. No additional eligible studies were identified. Of the 103 flagged abstracts, 99 were false positives, corresponding to a specificity of 99.8%.

### Study 5: APOC3 Antisense Oligonucleotides in Hypertriglyceridemia

The fifth validation study assessed APOC3-targeting antisense oligonucleotides for the treatment of hypertriglyceridemia. Four recently published meta-analyses served as benchmarks for this evaluation (49–52). A PubMed search yielded a total of 1,226 abstracts. Synthesa AI screened this corpus and flagged 37 abstracts for human review. Among these, the tool correctly identified all 10 RCTs previously included in the benchmark meta-analyses, corresponding to a sensitivity of 100%. Additionally, the tool identified one further eligible RCT that was published after the search cutoffs of the reference meta-analyses and thus not included in their final datasets (53). Of the 37 flagged abstracts, 24 were ultimately classified as false positives, resulting in a specificity of 98%.

### Study 6: Dexmedetomidine and Postoperative Delirium in Cardiac Surgery

The sixth validation study assessed Synthesa AI’s performance in replicating a meta-analysis by Hunt et al., which investigated the impact of perioperative dexmedetomidine administration on postoperative delirium in patients undergoing cardiac surgery (54). A combined search of PubMed and Scopus yielded 23,667 abstracts. Synthesa AI processed the entire corpus and flagged 235 abstracts for further review. All 12 benchmark RCTs were correctly identified by the tool, achieving a sensitivity of 100%. Importantly, no additional eligible studies were identified beyond those already included in the benchmark meta-analysis. Of the 235 flagged abstracts, 223 were classified as false positives, resulting in a specificity of 99.06%.

### Study 7: Coadministration of Pneumococcal Vaccines with Influenza or SARS-CoV-2 Vaccines

The seventh validation study examined the safety and immunogenicity of coadministration strategies involving pneumococcal vaccines with either influenza or SARS-CoV-2 vaccines. The benchmark reference was a meta-analysis by Rahimi et al. (55), which included 17 relevant studies. A highly sensitive PubMed search was conducted for this study, yielding a total of 69,207 abstracts. Synthesa AI screened the entire corpus and flagged 726 abstracts for manual review. All 17 benchmark studies were successfully identified by the tool , corresponding to a 100% sensitivity. Additionally, the tool identified two eligible studies published after the cutoff date of the reference meta-analysis (56, 57), as well as one relevant study that had been published earlier but was not included in the benchmark dataset (58). These findings increased the total number of relevant studies to 20. Of the 726 flagged abstracts, 703 were adjudicated as false positives, yielding a specificity of 99%.

### Study 8: Nebulized Antibiotics for Prevention of Ventilator-Associated Pneumonia (VAP)

The eighth validation study assessed Synthesa AI’s performance in replicating the results of a meta-analysis that examined the efficacy of nebulized antibiotics for preventing ventilator-associated pneumonia in critically ill patients (59). A total of 19,303 abstracts were retrieved from PubMed using a broad search strategy. Synthesa AI flagged 31 abstracts for manual adjudication. The tool correctly identified all four benchmark studies, achieving a sensitivity of 100%. No additional eligible studies were identified beyond those previously reported in the reference meta-analysis. Of the 31 flagged abstracts, 27 were deemed false positives, resulting in a specificity of 99.9%.

### Study 9: Vitamin D Supplementation for Prevention of Acute Respiratory Infections

The ninth validation study addressed the use of vitamin D supplementation to prevent acute respiratory infections, using a recently published meta-analysis by Jolliffe et al. as the benchmark reference (60). A total of 29,307 abstracts were retrieved via PubMed. Synthesa AI screened the entire dataset and flagged 269 abstracts for adjudication. All 46 benchmark studies were correctly identified by the tool, resulting in a sensitivity of 100%. In addition, Synthesa AI identified three additional RCTs that met inclusion criteria (61–63). These findings increased the total number of relevant studies to 49. Of the 269 flagged abstracts, 223 were ultimately excluded, resulting in a specificity of 99.3%.

### Cross-Study Performance Metrics

Across the nine validation studies conducted, Synthesa AI screened a total of 270,626 abstracts, a screening volume that represents a 1,380% increase over the 19,595 abstracts collectively reviewed in the 17 published benchmark meta-analyses used as reference standards. Despite the vast increase in scale, the tool flagged only 1,797 abstracts for manual review—representing a 91.7% reduction in screening burden— while maintaining perfect sensitivity (100%) across all studies. That is, Synthesa AI successfully identified every study included in the respective benchmark analyses, regardless of clinical domain, study design heterogeneity, or corpus size (**Table 1**).

**Table 1:** Summary of Synthesa AI’s performance metrics

| Topic | Bench<br>mark<br>Report<br>s | Number of<br>abstracts<br>screened | Number of<br>abstracts screened<br>in benchmark<br>reviews | Number of<br>abstracts<br>flagged for<br>human review | Number of<br>relevant studies<br>identified by<br>humans | Number of<br>studies<br>identified by<br>tool | Number of<br>additional<br>studies<br>identified by tool | Sen<br>siti<br>vity | Spe<br>cifi<br>city |
| --- | --- | --- | --- | --- | --- | --- | --- | --- | --- |
| APOC3<br>antisense<br>oligonucleotides | 4 | 1226 | 1565 | 24 | 10 | 11 | 1 | 100<br>% | 98<br>% |
| STEMI and<br>multivessel PCI<br>strategies | 6 | 5460 | 7199 | 162 | 24 | 26 | 2 | 100<br>% | 97.<br>20<br>% |
| Rescue inhalers<br>for asthma | 1 | 30719 | 4386 | 247 | 23 | 38 | 15 | 100<br>% | 99.<br>20<br>% |
| Antidepressants<br>for IBS | 1 | 43487 | 784 | 86 | 20 | 28 | 8 | 100<br>% | 99.<br>80<br>% |
| Dexmedetomidine and<br>postoperative<br>delirium | 1 | 23667 | 315 | 223 | 12 | 12 | 0 | 100<br>% | 99.<br>06<br>% |
| Pneumococcal<br>vaccine<br>coadministration | 1 | 69207 | 358 | 706 | 20 | 23 | 3 | 100<br>% | 99<br>% |
| Antiplatelet vs<br>anticoagulation<br>in HF | 1 | 48250 | 1647 | 99 | 4 | 4 | 0 | 100<br>% | 99.<br>80<br>% |
| Nebulized<br>antibiotics for<br>VAP | 1 | 19303 | 2441 | 27 | 4 | 4 | 0 | 100<br>% | 99.<br>90<br>% |
| Vitamin D for<br>acute<br>respiratory<br>infections | 1 | 29307 | 900 | 223 | 46 | 49 | 3 | 100<br>% | 99.<br>30<br>% |
| Totals | 17 | 270626 | 19595 | 1797 | 163 | 195 | 32 | 100<br>% | 99.<br>40<br>% |

Specificity across all validation exercises remained consistently high. The pooled specificity, defined as the proportion of correctly excluded abstracts among all those not included in benchmark reviews, was calculated at 99.4% (**Figure 1**). This estimate was derived from individual study-specific specificity rates, which ranged from 97.2% to 99.9%, and reflects the tool’s ability to minimize false positive rates even in extremely large and diverse literature corpora. Importantly, Synthesa AI identified a total of 32 relevant studies that were not included in the original benchmark publications. These additional studies were identified within the same pool of 1,797 abstracts flagged for human review, resulting in a 19.6% increase in relevant study yield relative to the 163 benchmark-included trials.

**Figure 1:**
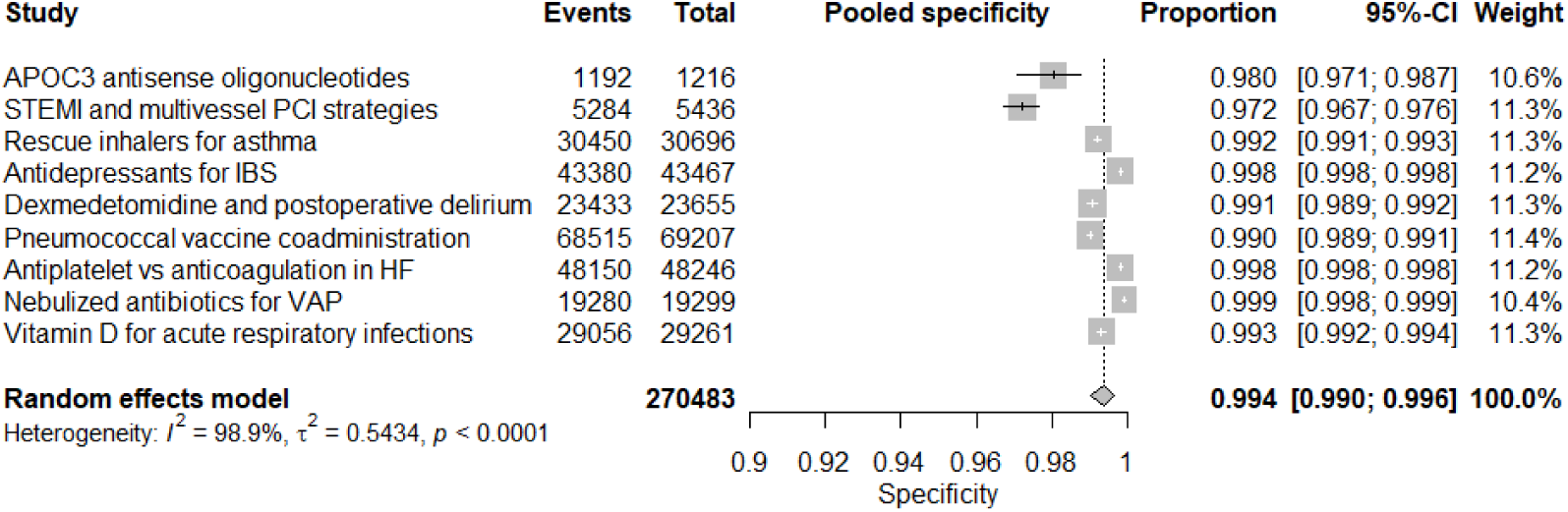
Synthesa AI’s pooled specificity across the nine validation studies.

## Discussion

This validation study provides compelling evidence that Synthesa AI, an LLM-based abstract screening tool, achieves performance metrics that meet or exceed those of traditional human screening in systematic reviews. Across nine independent validation exercises, Synthesa AI achieved perfect sensitivity (100%) in identifying all relevant studies included in benchmark meta-analyses, while maintaining a pooled specificity of 99.4%. These findings demonstrate not only the tool’s capacity to emulate expert-level screening behavior but also its operational feasibility for large-scale deployment in evidence synthesis workflows.

A particularly salient finding of this evaluation is Synthesa AI’s capacity to substantially reduce the manual workload associated with abstract screening. Across a cumulative corpus of over 270,000 abstracts, the tool flagged only 1,797 abstracts for human review, corresponding to a 91.7% reduction in reviewer burden compared to full manual screening. Importantly, this screening efficiency did not come at the cost of reduced sensitivity or precision. On the contrary, the tool identified 32 additional relevant studies beyond those included in the 17 benchmark meta-analyses, a 19.6% increase in yield.

The clinical and methodological implications of these findings are substantial. First, the study indirectly quantifies the limitations of human-led abstract screening, which is inherently constrained by fatigue, inter-reviewer variability, and practical resource limitations. The observation that approximately one in five relevant studies were absent from benchmark reviews—despite their adherence to stated inclusion criteria—raises important concerns regarding the completeness and reproducibility of traditional systematic reviews. Synthesa AI, in contrast, demonstrated perfect reproducibility across all validation runs; its deterministic prompt architecture ensures that identical inputs yield identical outputs, a property not shared by either human reviewers or probabilistic machine learning classifiers.

Second, the findings support the viability of integrating LLM-based screening tools into regulatory and academic settings where methodological rigor, auditability, and transparency are non-negotiable. Synthesa AI’s architecture enables full traceability of every inclusion and exclusion decision, with rationales and flagged terms available for documentation and adjudication. This feature aligns well with emerging frameworks such as the U.S. Food and Drug Administration’s Good Machine Learning Practice (GMLP) guidelines, which emphasize transparency, reproducibility, and human oversight in AI-assisted decision-making (64). As such, Synthesa AI holds promise not only as an academic research instrument but also as a tool for regulatory-grade literature reviews in contexts such as pharmacovigilance, clinical evaluation reports, and health technology assessments.

Third, the results of this study reinvigorate the long-standing vision of living systematic reviews. By enabling full corpus screening in near-real time, Synthesa AI renders feasible what had previously been considered operationally prohibitive—namely, the continuous updating of systematic reviews in response to newly published evidence. This capability could transform the responsiveness of evidence synthesis to fast-moving therapeutic domains such as infectious disease, oncology, and cardiovascular medicine, where the evidence base evolves rapidly and policy decisions must keep pace.

Despite its strengths, the present study is not without limitations. The calibration of Synthesa AI was based on known included studies in the benchmark reviews, which may introduce bias by optimizing the tool for sensitivity against those specific inclusion patterns. This means that generalizability to entirely novel topics without an existing gold standard cannot be inferred directly from the current results. Future work will expand on this approach to further evaluate the tool’s de novo screening performance under real-world, prospective conditions.

In conclusion, Synthesa AI delivers a transformative improvement in the accuracy, completeness, and efficiency of abstract screening for systematic reviews. By substantially reducing reviewer burden, identifying previously missed studies, and enabling reproducible decision-making at scale, Synthesa AI addresses longstanding challenges in evidence synthesis. These findings set a new benchmark for what can be achieved in systematic review methodology and point toward a future in which LLM-based tools play a central role in the curation of clinical evidence.

Figure Legend: Forest plot of pooled specificity across the 9 topics

## Data Availability

All data produced in the present study are available upon reasonable request to the authors

